# SARS-CoV-2 exposure, symptoms and seroprevalence in health care workers

**DOI:** 10.1101/2020.06.22.20137646

**Authors:** Ann-Sofie Rudberg, Sebastian Havervall, Anna Månberg, August Jernbom Falk, Katherina Aguilera, Henry Ng, Lena Gabrielsson, Ann-Christine Salomonsson, Leo Hanke, Benjamin Murrell, Gerald McInerney, Jennie Olofsson, Eni Andersson, Cecilia Hellström, Shaghayegh Bayati, Sofia Bergström, Elisa Pin, Ronald Sjöberg, Hanna Tegel, My Hedhammar, Mia Phillipson, Peter Nilsson, Sophia Hober, Charlotte Thålin

## Abstract

**Background:** SARS-CoV-2 may pose an occupational health risk to health care workers, but the prevalence of infections in this population is unknown. We examined the seroprevalence of SARS-CoV-2 antibodies among health care workers at a large acute care hospital in Stockholm, Sweden. We determined correlations between seroprevalence, self-reported symptoms and occupational exposure to SARS-CoV-2.

**Methods and findings:** All employees at Danderyd Hospital (n=4375) were invited to participate in a cross-sectional study. 2149 employees from all hospital departments were enrolled in the study between April 14^th^ and May 8^th^ 2020. Study participants completed a questionnaire consisting of symptoms compatible with SARS-CoV-2 infection since January 2020 and occupational exposure to patients infected with SARS-CoV-2. IgG antibodies against SARS-CoV-2 were analyzed using a multiplex assay evaluated to have 99.4% sensitivity and 99.1% specificity. The over-all seroprevalence among 2149 participants was 19.1% (n=410). There was no difference in age or sex between seropositive and seronegative participants. The symptoms with the strongest correlation to seroprevalence were anosmia and ageusia, with odds ratios of 28.4 (p=2.02*10^-120) and 19.2 (p=1.67*10^-99) respectively. Seroprevalence was strongly associated with patient-related work (OR 2.9, p=4.24*10^-8), covid-19 patient contact (OR 1.43, p=0.003), and occupation as assisting nurse (OR 3.67, p=2.16*10^-9).

**Conclusion:** These results demonstrate that anosmia and ageusia should be included in screening guidance and in the recommendations of self-isolation to reduce further spread of SARS-CoV-2. The results furthermore imply an occupational health risk for SARS-CoV-2 infection among hospital workers. Continued measures are warranted to assure healthcare worker safety and reduce transmission from health care settings to the community during the covid-19 outbreak.

## Introduction

The novel corona virus coined severe acute respiratory syndrome coronavirus-2 (SARS-CoV-2), causes the coronavirus disease 2019 (COVID-19). The World Health Organization (WHO) declared Covid-19 to be a pandemic on March 11, 2020 (1) and currently more than 8 million cases have been reported, leading to over 440 000 deaths. The first laboratory confirmed case of covid-19 infection in Sweden was observed 31^st^ January 2020, escalating to 22317 cases and more than 5000 deaths as of June 17^th^ 2020 (2). Epidemiology of the covid-19 infection is, however, largely based on cases requiring hospitalization, and little is known about the true extent of the disease. Serological population-based investigations provide a useful tool in the estimation of the number of individuals who have been infected with the SARS-CoV-2 virus and who may be at reduced risk for re-infection. Several estimations of population-based seroprevalence are emerging, ranging from 4.4% in France (3), 4.6 % in Los Angeles (4), and 7.3% in Stockholm (2), all from April-May 2020.

The Swedish main objective with the actions coupled to the covid-19 pandemic has been similar to that of most other countries; to reduce the spread of the infection. However, instead of a full lock down, the strategy has been to keep parts of the society open. Events where more than 50 people take part have been banned, but the majority of actions to reduce the spread have relied upon voluntary compliance with the Public Health Authority’s evolving set of recommendations (2). These recommendations are non-compulsory but individuals are expected to follow them, despite the lack of fines for any failure. The inhabitants have been urged to work from home if possible and to avoid travels. Further, limited social contacts are encouraged, specifically among individuals with greatest risk for covid-19 complications is clearly stated. In contrast to many other countries, however, preschools and grade schools have remained open.

Health care workers (HCW) are exposed to the virus at a greater extent than society as a whole, and may be considered at an elevated risk of infection. During the previous SARS epidemic, HCW comprised more than 20% of all cases (5-7), and reports of high prevalence of SARS-CoV-2 infection amongst HCW are now emerging (8, 9). This raises concerns about the safety of front-line HCW, and the risk of healthcare system collapse as well as transmission from health care settings to the community. Little is, however, known about the occupational risk of HCW to SARS-CoV-2 infection. We therefore determined the SARS-CoV-2 antibody prevalence among HCW with occupational exposure to patients with and without covid-19. Through the mapping of self-reported symptoms, we furthermore identified symptoms most predictive of SARS-CoV-2 infection.

## Material and Methods

### Hospital setting

This cross-sectional study was conducted at Danderyd Hospital, an acute-care 530-bed hospital providing both general and specialized hospital care. With a catchment area of 600 000 individuals, a total of 722 patients with confirmed covid-19 had been admitted to the hospital during the study period. The hospital guidelines require PPE to be worn by all employees with direct contact with confirmed or suspected SARS-CoV-2 infected patients. Face shield, FFP3/FFP2/N95 mask and sleeveless plastic apron was recommended in general wards during aerosol generating procedures (AGPs), and at all times in ICU. Aerosol filtering face mask is replaced by surgical face mask IIR during non-AGP. During the study period, reverse-transcriptase polymerase chain reaction (RT-PCR) viral RNA detection of nasopharyngeal or oropharyngeal swabs was not available for hospital employees.

### Study population

All employees at Danderyd Hospital (n=4375) were invited by e-mail and through information on the hospital intranet to participate in the study. Consecutive study inclusion took place between 15^th^ of April to 8^th^ of May 2020. Participants were eligible to participate in the study irrespective of whether they had had symptoms since the covid-19 outbreak onset or not. The only exclusion criterion was absence from work due to illness. Informed consent, study inclusion and appointment for blood sampling were obtained using a smartphone-based app and verification through electronic identification and signature. Participants completed a questionnaire comprising demographics (age and sex), self-reported predefined symptoms compatible with covid-19 since 1^st^ of January 2020, occupation, work location and self-reported exposure to patients infected with covid-19 prior to blood sampling. The questionnaire consisted of eleven pre-defined symptoms (fever, headache, anosmia, ageusia, cough, malaise, common cold, abdominal pain, sore throat, shortness of breath) including the alternative “no ongoing or prior symptoms since January 1^st^ 2020”. Participants were asked to grade the symptoms as mild or severe, and to document symptom onset. Participants were asked to state their occupation as physician, nurse, assisting nurse or other health care personnel, as well as department of employment in free text. Due to frequent relocation of hospital staff during the pandemic, participants were further asked to state whether they had direct patient contact and whether they had worked in covid-19 departments, (covid-19 zone of the emergency department, covid-19 transit ward (admitting patients awaiting laboratory confirmation of covid-19 infection), covid-19 general wards and covid-19 intermediate and intensive care units IMU/ICU). The portion of study participants answering all questions was 96.3%.

### Serological analyses of antibodies

Plasma samples were prepared from whole blood following centrifugation for 20 min at 2000 g at room temperature, viral inactivation for 30 minutes in 56 °C, and stored at −80°C until further analyses. Serological analyses of IgG were performed. IgG reactivity was measured towards four different virus protein variants (Spike trimers comprising the prefusion-stabilized spike glycoprotein ectodomain (10) (in-house produced, expressed in HEK and purified using a C-terminal Strep II tag) Spike S1 domain (Sino Biological, expressed in HEK and purified using a C-terminal His-tag), Spike Receptor Binding Domain (RBD) (in-house produced, expressed in HEK and purified using C-terminal His-tag) and Nucleocapsid protein (Acro Biosystems, expressed in HEK and purified using a C-terminal His-tag) and analyzed using a multiplex antigen bead array in high throughput 384-plates format using a FlexMap3D (Luminex Corp) (11). The cutoff values used in the study was chosen after analyzing 154 positive samples (defined as PCR-positive individuals with mild to severe symptoms sampled more than 14 days after onset) and 321 negative samples (defined as samples collected 2019 and earlier, including confirmed infections with non-SARS-CoV-2 corona viruses). Based on these control samples, the method was calculated to have 99.4% sensitivity and 99.1% specificity. Plasma IgG were captured on the antigen coated beads and detected by fluorescent anti-hIgG. The read out consisted of the bead-based median fluorescent intensity (MFI) and count of number of beads for each antigen (bead ID) in each sample. To be assigned as an IgG positive plasma sample, reactivity against at least two of the four different variants of the viral antigens was demanded and the cutoff was defined as signals above the mean+6 SD of the 12 negative controls included in each analysis.

### Statistical analyses

Descriptive analyses were made on baseline characteristics and the number of observations, presented as number and percentages. Categorical variables are presented as proportions, compared with the Fisher’s exact test and reported as odds ratio (OR) and 95% confidence interval (CI). Continuous variables are presented as means with standard deviations (SD) and compared with the unpaired t-test. The group of HCW with patient-related work (n=1764) was compared to the group of HCW with non-patient related work (n=305). The group of HCW with patient related work was further divided into covid-19 patient contact and non-covid-19 patient contact, and these two sub groups were compared to each other. Within the group of HCW with patient related work, occupations as assisting nurses (n=428), nurses (n=636), other health-care personnel (n=254), and physicians (n=439) were compared to the group with non-patient-related work. Statistical analyses and visualizations were performed in R, using packages tidyverse, lubridate, rlang, pander, knitr, and UpSetR (RStudio Team 2019, Boston, USA).

The study complied with the declaration of Helsinki, and informed consent was obtained on all participants. The study protocol was approved by the Stockholm Ethical Review Board (dnr 2020-01653). This report conforms to the STROBE guidelines.

## Results

A total of 2149 HCW were included in the study. The majority of study participants were women (85%) and the mean age was 44 (SD 12) years. Patient contact was reported by 1764 participants (85%), and 962 (46%) participants reported covid-19 patient contact. The group with patient contact comprised 439 (25%) physicians, 636 (36%) nurses, 428 (24%) assisting nurses, and 254 (14,5%) other healthcare staff (occupation was missing in 0.5%). (Table 1).

**Table 1.** Demographics, symptomatology and occupational exposure of seropositive and seronegative study participants.

|  | All<br>n=2149 | Seronegative<br>n=1739 | Seropositive<br>n=410 | Missing<br>(all) | Missing<br>(Seronegative) | Missing<br>(Seropositive) |
| --- | --- | --- | --- | --- | --- | --- |
| Age (years), mean (SD) | 44(12) | 44(12) | 43(12) | 2 | 1 | 1 |
| Female, n (%) | 1815(85) | 1475(85) | 340(83) | 3 | 3 | 0 |
| Male, n (%) | 331(15) | 261(15) | 70 (17) | 3 | 3 | 0 |
| Symptoms since 1st January 2020, n (%): |  |  |  |  |  |  |
| Fever | 538(25) | 304(17) | 234(57) | 0 | 0 | 0 |
| Headache | 991(46) | 722(42) | 269(66) | 0 | 0 | 0 |
| Anosmia | 283(13) | 66(4) | 217(53) | 0 | 0 | 0 |
| Ageusia | 289(13) | 85(5) | 204(50) | 0 | 0 | 0 |
| Cough | 716(33) | 503(29) | 213(52) | 0 | 0 | 0 |
| Malaise | 912(42) | 644(37) | 268(65) | 0 | 0 | 0 |
| Common cold | 738(34) | 557(32) | 181(44) | 0 | 0 | 0 |
| Abdominal symptoms | 382(18) | 261(15) | 121(30) | 0 | 0 | 0 |
| Sore throat | 822(38) | 660(38) | 162(40) | 0 | 0 | 0 |
| Shortness of breath | 303(14) | 205(12) | 98(24) | 0 | 0 | 0 |
| No symptoms | 460(21) | 423(24) | 37(9) | 0 | 0 | 0 |
| Degree of symptoms, n (%): |  |  |  |  | 0 | 0 |
| Mild | 1573(73) | 1253(72) | 320(78) | 0 | 0 | 0 |
| Severe | 116(5) | 63(4) | 53(13) | 0 | 0 | 0 |
| Exposure, n (%): |  |  |  |  |  |  |
| Physicians | 439(21) | 355(21) | 84(21) | 80(4) | 67(4) | 13(3) |
| Nurses | 636(31) | 497(30) | 139(35) | 80(4) | 67(4) | 13(3) |
| Assisting nurses | 428(21) | 319(19) | 109(27) | 80(4) | 67(4) | 13(3) |
| Other healthcare staff | 254(12) | 220(13) | 34(9) | 80(4) | 67(4) | 13(3) |
| Employees with no patient contact | 305(15) | 279(17) | 26(7) | 80(4) | 67(4) | 13(3) |
| Direct patient contact | 1764 (85) | 1393 (83) | 371 (93) | 80(4) | 67(4) | 13(3) |
| Direct contact with covid patients | 962 (46) | 734 (44) | 228 (57) | 80(4) | 67(4) | 13(3) |

Overall, 410 study participants (19.1%) were seropositive for IgG. There was no difference in age or sex between seropositive and seronegative individuals. (Table 1).

Interestingly, among seropositive individuals, 37 individuals (9%) reported no symptoms at all, 320 individuals (78%) reported mild symptoms, and only 53 individuals (13%) reported severe symptoms. The most frequently reported symptoms in this group were headache (66%), malaise (65%), fever (57%), anosmia (53%), cough (52%) and ageusia (50%), while abdominal pain (30%) and dyspnea (24%) were the least reported symptoms (Figure 1).

**Fig 1.**
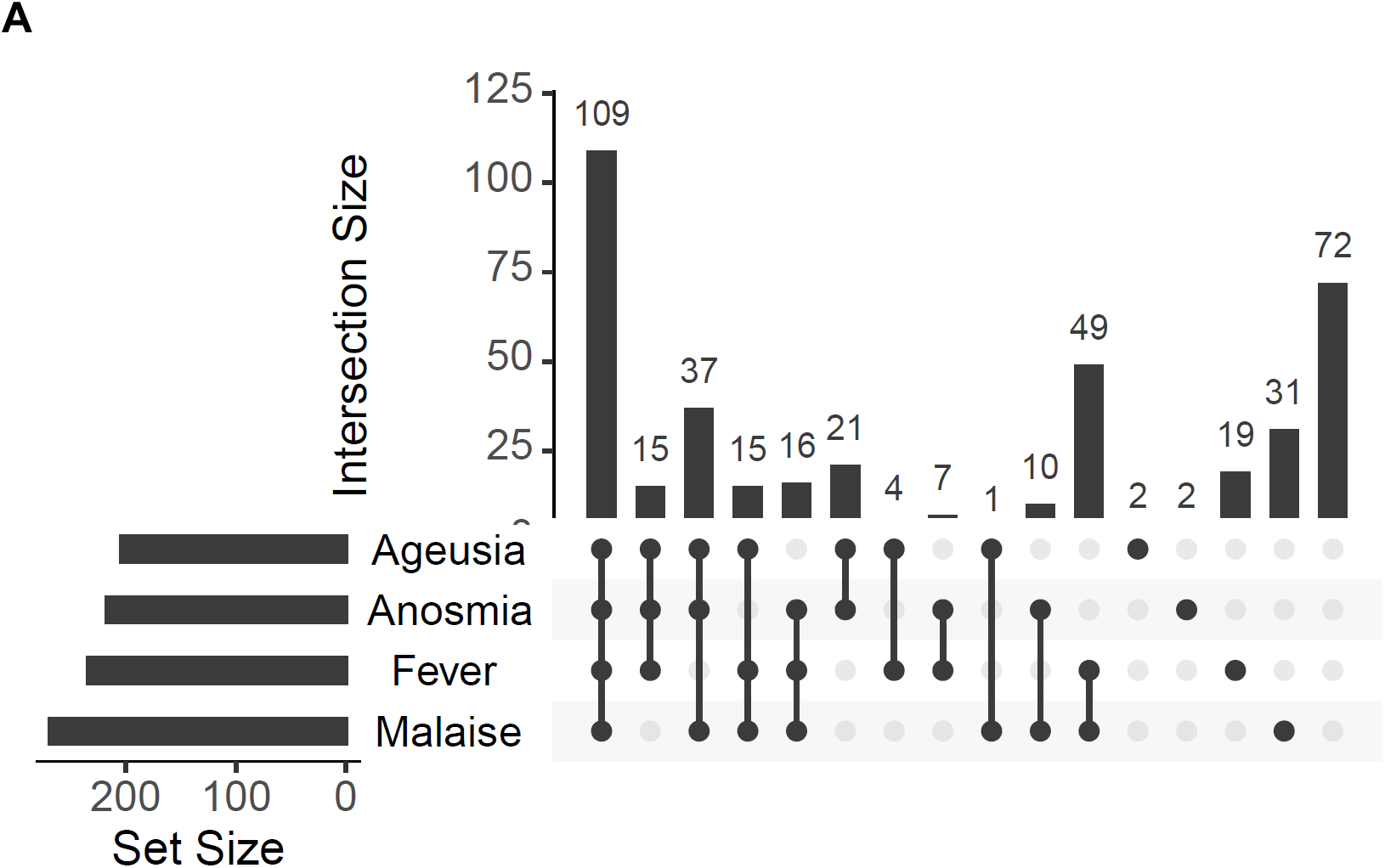
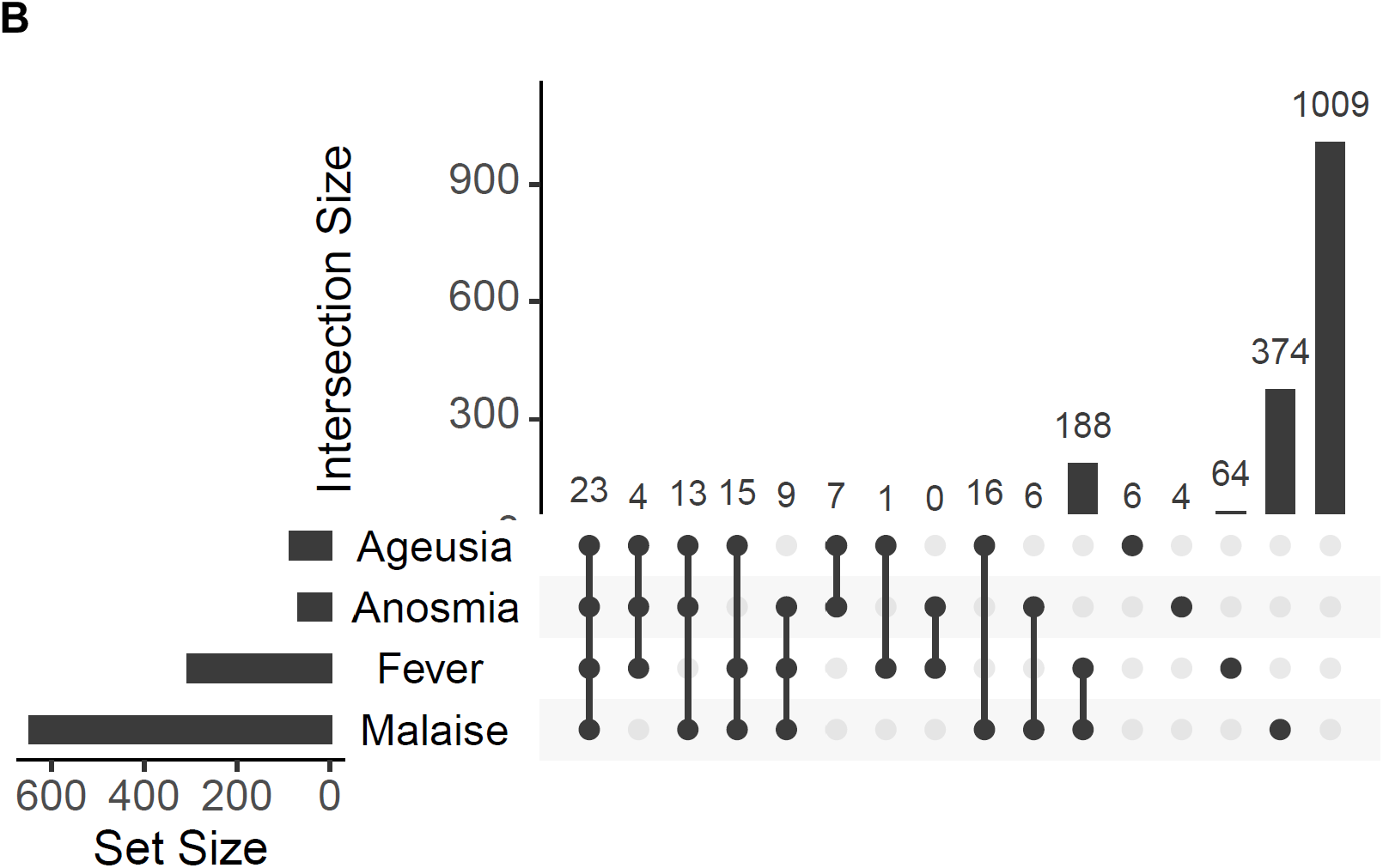
Symptomatology in seropositive (A) and seronegative (B) individuals. Horizontal bars to the left represent the total number of participants in each group reporting the specifically denoted symptom. Vertical bars show the total number of participants in each group reporting symptoms symbolized with black dot(s) in the corresponding column.

Symptoms with the strongest association to seroprevalence were anosmia (OR 28.43; p=2.02*10^-120), ageusia (OR 19.21; p=1.67*10^-99) and fever (OR 6.27; p=4.24*10^-8). The only symptom that did not differ in prevalence between seronegative and seropositive participants was sore throat (OR 1.07; p=0.572). Two symptom triads were strongly associated with seroprevalence; anosmia and/or ageusia, malaise and fever (OR 18.62, p=1.64*10^-69), as well as anosmia and/or ageusia, malaise and cough (OR 11.90, p=2.01*10^-49). (Figure 2). The positive predictive value (PPV) and negative predictive value (NPV) of any three of anosmia, ageusia, malaise and fever were 0.75 and 0.88 respectively.

**Fig 2.**
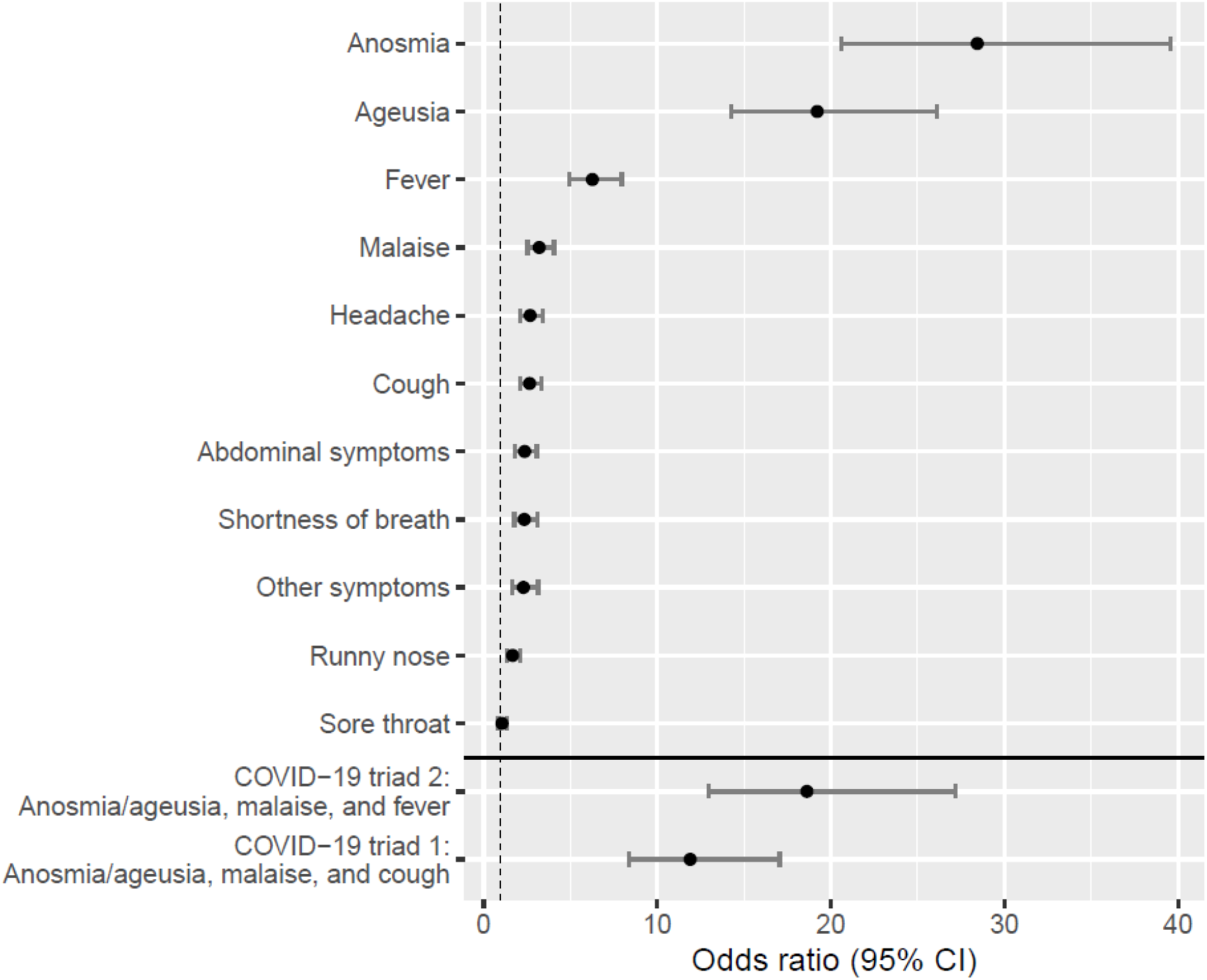
Associations between prior symptoms and seroprevalence of SARS-CoV 2 IgG antibodies. The figure shows odds ratios of seropositivity for individually reported symptoms.

Seroprevalence was associated to patient-related work, with 21% among 1764 study participants with patient contact vs. 9% among 305 study participants without patient contact (OR 2.9, p=4.24*10^-8). Seroprevalence was also significantly higher among study participants with covid-19 patient contact than among study participants without known exposure to covid-19 patients; 24% vs. 18% (OR 1.43, p=0.003), (Figure 3).

**Fig 3.**
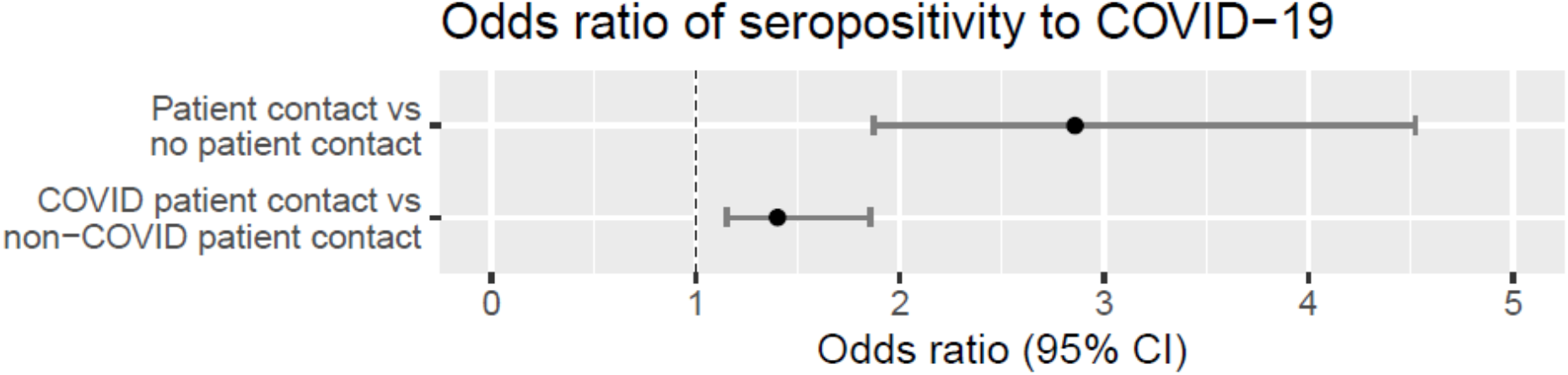
Association between occupational exposure and seroprevalence of SARS-CoV 2 IgG antibodies. The figure shows odds ratios of seropositivity if patient contact and if covid19 patient contact.

However, the seroprevalence among study participants with patient contact but without known covid-19 contact remained higher than that found among study participants without patient contact (OR 2.33, p<0.0001). The association between patient contact and seroprevalence remained significant regardless of whether the occupation was physician (OR 2.54, p=5.69*10^-5), nurse (OR 3.0, p=2.13*10^-7), or assisting nurse (OR 3.67, p=2.16*10^-9), (Figure 4), but the seroprevalence was higher among assisting nurses (25%) and nurses (22%) than among physicians (19%) and other medical staff (13%).

**Fig 4.**
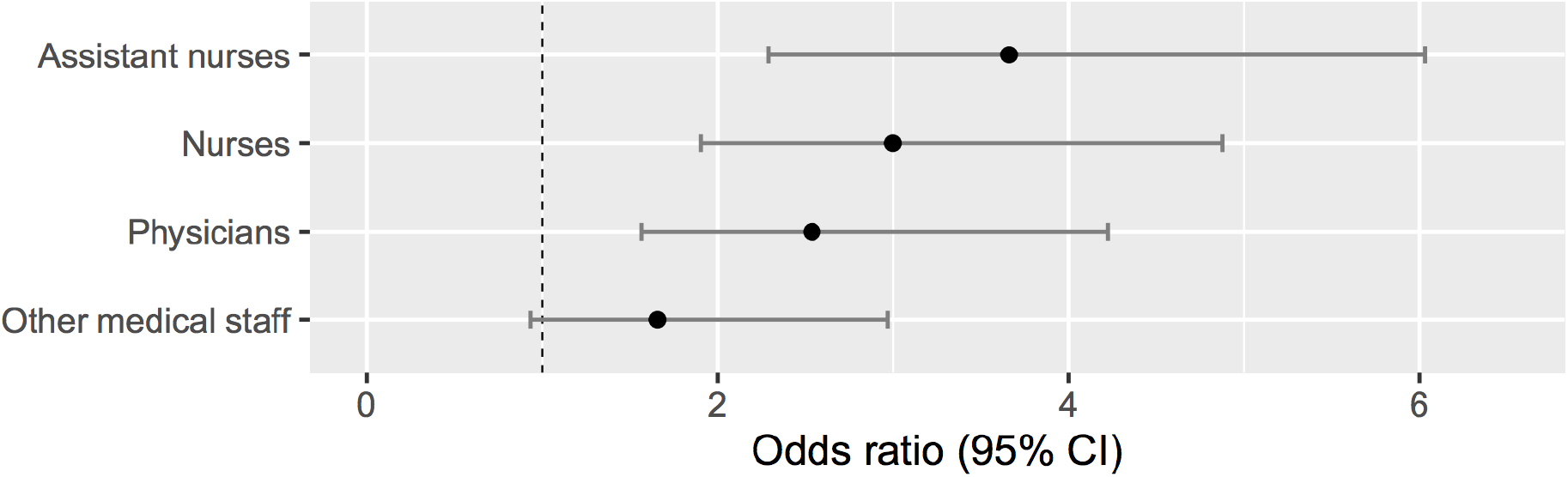
Associations between occupations and seroprevalence of SARS-Cov 2 IgG antibodies among HCW with patient contact. The figure shows odds ratios of seropositivity and occupation within the group of HCW with patient contact.

## Discussion

This is, to our knowledge, the first large serological investigation supporting an occupational risk of SARS-CoV-2 transmission to HCW. Among the enrolled 2149 study participants, one in five were seropositive, suggesting prior or still on-going infection with SARS-CoV-2. The seroprevalence was significantly associated to both patient-related work and covid-19 patient contact, and was substantially higher than what was reported in the general population of Stockholm (7.3%) during the same time period and using the same serological test (2), suggesting an occupational health risk among HCW. We furthermore report symptoms strongly associated with SARS-CoV-2 infection, which may aid in health care personnel screening guidance and in the recommendations of self-isolation.

The portion of seropositive study participants reporting no prior symptoms in this cohort is lower than several outbreak investigations suggesting that up to 50% of covid-19 infections remain subclinical (12). However, the vast majority of study participants reported mild symptoms that are difficult to distinguish from other respiratory infections. Identification of symptoms predictive of SARS-CoV-2 infection, single or in combination, is essential for the quest of screening guidance and in the recommendations of self-isolation to prevent further spread. In a large population-based study including 9 282 HCW in the Unites States, 92% reported having at least one symptom among fever, cough, and dyspnea (13), and this symptom triad has been reported hallmark symptoms of covid-19 infection (14-16). Current covid-19 HCW screening guidance from the Centers for Disease Control and Prevention (17) therefore includes assessing fever, cough and dyspnea. Although fever was one of the most common symptoms among the seropositive HCW in this study, anosmia and ageusia were the symptoms most predictive of SARS-CoV-2 infection. Of these, anosmia is emerging as a key symptom in covid-19 (18, 19), and our results are in line with the recent large multinational population-based cohort study investigating potential symptoms of covid-19, reporting a strong association between anosmia, ageusia and covid-19 (19). We furthermore report a strong association between the symptom combination anosmia and/or ageusia, malaise and fever and seroprevalence, suggesting that these symptoms may be included in routine screening guidance.

The occupational risk of HCW to be infected with SARS is well documented (20-22). Emerging reports now highlight the risk of occupational transmission also of SARS-CoV-2 (9, 13, 22-28), and contamination of SARS-CoV-2 RNA has been demonstrated to be widespread across hospital environmental surfaces (29). The use of proper PPE has been prioritized since the start of the covid-19 outbreak (24, 30), but our results support the early reports of patient care being a risk factor for SARS-CoV-2 infection. When comparing the seroprevalence among study participants with non-patient-related work with those with patient-related work, the discrepancy was large (9% vs 21%), and the seroprevalence of 9% among the study participants with non-patient-related work was in line with the at the time estimated seroprevalence of 7.3% in Stockholm. The seroprevalence was also higher among study participants with covid-19 patient contact than among study participants with patient contact but without known contact with covid-19 patients, reinforcing a patient-related transmission of SARS-CoV-2 to HCW. Notably, the seroprevalence among study participants with patient contact but without known contact with covid-19 patients remained elevated compared with the group of study participants without any patient contact at all. The reason for this is unclear, but difficulties in distinguishing covid-19 patients from patients not infected with SARS-CoV-2 may have contributed to a transmission to HCW in non-covid-19 departments. The high seroprevalence among assistant nurses and nurses further supports transmission from patients to HCW considering that these occupations involve the most patient-near contact.

The results observed in this study are strengthened by the large sample size of individuals representing all departments at Danderyd Hospital. Furthermore, the response rate of self-reported data was close to 100%. The self-reporting of occupational exposure to SARS-CoV-2 was found to be superior to occupational location in the hospital data base since a large portion of study participants are re-located during the covid-19 pandemic. The results are further strengthened by the use of a highly sensitive and specific laboratory-based antibody assay. The method is currently being evaluated by the use of almost 500 reference samples with a high sensitivity and specificity. This is in contrast with many other serological test methods, where the sample set used for evaluation is very low and thereby also the certainty of the test method (31).

The study also has several limitations worth noting. Seroprevalence is dynamic, and the data we present in this cohort represents the prevalence of prior or still on-going infection in April-May 2020. A new cross-sectional investigation will yield different results. Another limitation lies in the nature of self-reported data, yielding a risk of recall bias. Anosmia and ageusia have been widely pointed out as potential covid-19 symptoms in media, which may have influenced responses. Participants were asked to document symptoms over the prior 3-4 months, and it is not certain that reported symptoms were caused by SARS-CoV-2 infection in seropositive participants, who may have been infected by other respiratory viruses in addition to SARS-CoV-2 in the months prior to study inclusion. The options of occupation, patient contact and exposure to covid-19 patients were, however, objective variables. Study participation was voluntary, and a selection bias cannot be excluded. Symptomatic individuals may have been more likely to participate, and employees not at work due to illness during the study period were excluded, which could have influenced the over-all seroprevalence. However, possible selection bias would apply to all sub groups, and comparisons between groups were likely unaffected. Appointments for study inclusion were furthermore released continuously at diurnal random times throughout the study period, ensuring that various occupations, regardless of work schedule, were able to participate. Finally, although a clear association was observed between seroprevalence, patient contact, covid-19 patient contact and occupation, we cannot rule out possible cluster spread among study participants regardless of occupational exposure to SARS-CoV-2.

In conclusion, anosmia and ageusia are common symptoms in SARS-CoV-2 infection, and should be considered, together with fever, when guiding screening and in the recommendations of self-isolation. HCW with direct patient contact are at increased risk of SARS-CoV-2 infection, and continued and enhanced efforts to protect HCW and to reduce transmission from hospital to the community are warranted. These measures may limit the ongoing pandemic of SARS-CoV-2.

## Data Availability

Yes

## Acknowledgment

We are grateful to Carola Jonson, Sofie Lundin, Tina Johnsson, Camilla Redhevon, Mitra Samadi, Bibi Fundell, Ami Sjöblom, Sara Juhlin, Emelie Karlsson, Lina Petersson, Julia Alm Baker, Eva Edström, Frehiywet Tes, Anna Weimer, Mery Millberg at Danderyd Hospital for assisting in administration and blood sampling. We thank Richard Scholvin for technical support with the smart phone app and assisting with data information. We are grateful to David Just, Cornelia Westerberg, Mimmi Olofsson and Sara Mravinacova for technical assistance in performing and developing the serology assays. We are grateful to Karolinska University Laboratory and Public Health Agency of Sweden for providing negative and positive control samples. We also would like to thank Gunilla Karlsson Hedestam for fruitful discussions.

